# The magnitude and cross reactivity of SARS-CoV-2 specific antibody responses in vaccinated and unvaccinated Sri Lankan children and association with nutrition status

**DOI:** 10.1101/2023.12.18.23300176

**Authors:** Chandima Jeewandara, Maneshka Vindesh Karunananda, Suranga Fernando, Saubhagya Danasekara, Gamini Jayakody, S. Arulkumaran, N.Y. Samaraweera, Sarathchandra Kumarawansha, Subramaniyam Sivaganesh, P. Geethika Amarasinghe, Chintha Jayasinghe, Dilini Wijesekara, Manonath Bandara Marasinghe, Udari Mambulage, Helanka Wijayatilake, Kasun Senevirathne, A.D.P Bandara, C.P. Gallage, N.R. Colambage, A.A. Thilak Udayasiri, Tharaka Lokumarambage, Y. Upasena, W.P.K.P. Weerasooriya, seroprevalence study group, Tiong Kit Tan, Alain Townsend, Graham S. Ogg, Gathsaurie Neelika Malavige, Study groups, Lahiru Perera, Pradeep Pushpakumara, Laksiri Gomes, Jeewantha Jayamali, Inoka Sepali Aberathna, Thashmi Nimasha, Madushika Dissanayake, Shyrar Ramu, Deneshan Peranantharajah, Hashini Colambage, Rivindu Wickramanayake, Harshani Chathurangika, Farha Bary, Sathsara Yatiwelle, Michael Harvie, Maheli Deheragoda, Tibutius Jayadas, Shashini Ishara, Dinuka Ariyaratne, Shashika Dayarathna, Ruwanthi Wijekulasuriya, Chathura Ranathunga

**Author notes:** Contributed equally (Chandima Jeewandara, Maneshka Vindesh Karunananda, Suranga Fernando). Correspondence should be addressed to: Prof. Neelika Malavige DPhil (Oxon), FRCP (Lond), FRCPath (UK) Allergy Immunology and Cell Biology Unit, Department of Immunology and Molecular Medicine, Faculty of Medical Sciences, University of Sri Jayawardanapura, Sri Lanka.

## Abstract

**Background:** We investigated the seropositivity rates of Sri Lankan children in a large island wide serosurvey, to determine the magnitude and breadth of antibody responses to SARS-CoV-2 variants and the association with the vaccination and nutritional status to understand the likely impact of newer variants in Sri Lanka.

**Methods:** Using the WHO UNITY protocol, we recruited 5207 children, aged 10 to 20 years, representative of the 9 provinces of Sri Lanka, and assessed seropositive rates, ACE2 blocking antibodies and antibodies to BA.2.75 and XBB.1.5, in vaccinated and unvaccinated children. Anthropometric measurements were taken to determine the association between nutrition status and antibody levels.

**Results:** 3111/3119 (99.7%) vaccinated and 2008/2088 (96.2%) of unvaccinated children, were seropositive for SARS-CoV-2. 2984/3111 (95.9%) of vaccinated children had ACE2 blocking antibodies above the cut-off threshold, compared to 1346/2008 (67.0%) of unvaccinated children. 27.2 % unvaccinated children had positive antibody responses to BA.2.75 and 30.7% to XBB.1.5, while 64.3% of vaccinated had a positive response to BA.2.75 and 43.1% to XBB.1.5. Vaccinated children had significantly higher titres of total antibodies, ACE2 blocking antibodies and antibodies to XBB.1.5 and BA.2.75 than unvaccinated children. However, the vaccinated had significantly higher responses to BA.2.75 (p<0.0001), than XBB.1.5. Unvaccinated children, with <3rd BMI centile had significantly lower ACE2 blocking antibodies than other groups.

**Discussion:** The high seropositivity rates and antibody titres to SARS-CoV-2 variants in unvaccinated children, suggests that they are likely to have been infected more than once with SARS-CoV-2. The implications of lower antibody levels in undernourished children should be further investigated.

## Background

Despite high vaccination rates and high seroprevalence rates in many countries, outbreaks of COVID-19 still occur in many regions, with the XBB.1.5 sub-lineages of omicron EG.5.1, XBB.2.3 and XBB.1.16 dominating [1, 2]. Although the number of hospital admissions and case fatality rates are low, COVID-19 still causes a significant impact on health care systems in some countries [1]. Many high-income countries and some lower-middle income countries are administering booster doses to their populations with updated versions of the COVID-19 vaccines, incorporating the XBB.1.5 variant [3]. While many countries are making these updated COVID-19 booster doses only available to vulnerable individuals, the CDC in USA has recommended these vaccines to all individual above the age of six months [3].

Sri Lanka experienced many COVID-19 outbreaks in the past, with high mortality rates, especially during the outbreak due to the delta variant [4]. However, deaths have been predominantly among the adults, with children rarely developing severe disease, as seen in all other countries [5, 6]. Although 28.5% of the population in Sri Lanka are children (<18 years of age), they accounted for <18% of reported cases of COVID-19, possibly due to the asymptomatic nature of infection among children [7, 8]. Therefore, the reported number of cases are unlikely to reflect true infection rates among children and seroprevalence studies are required to understand the true infection rates in the population. Furthermore, unlike many other countries globally and in the region, Sri Lanka did not offer any bivalent booster doses for individuals in Sri Lanka and children >12 years of age were offered only two doses of the Pfizer BioNTech (BNT162b2) vaccine. Therefore, the Sri Lankan population were expected to be vulnerable to the sub-lineages of the omicron variant, especially the XBB.1.5 and its sub lineages, which are very immune evasive [9].

The nutritional status affects immunity to many viral infections, and the intake of micro and macronutrients has shown to affect susceptibility to SARS-CoV-2 infection [10]. Obesity has shown to be associated with a significantly lower antibody responses to COVID-19 vaccines [11, 12]. However, there are limited data on antibody responses to SARS-CoV-2 following natural infection or vaccination in those who are underweight or malnourished. As Sri Lanka is going through an economic crisis, it has been estimated that 28.2% of the population are below the poverty line (<US$ 3.65 per day) in 2023 with 2.9 million children in urgent need of humanitarian assistance [13]. Therefore, it would be important to determine if undernutrition affects the antibody levels to SARS-CoV-2 and the breadth of responses. This will enable us to understand the extent of population immunity that will affect transmission dynamics when novel variants are introduced into the population.

In this study, to understand the immunity to SARS-CoV-2 in the population and to investigate the magnitude and breadth of antibody responses in undernourished and with normal nutrition status, we carried out an island wide serosurvey to detect the presence of antibodies to SARS-CoV-2 using the WHO UNITY protocol and also assessed the neutralizing antibody responses (Nabs), using a surrogate neutralizing antibody assay and also assessed the antibody responses to currently circulating SARS-CoV-2 omicron sub-lineages.

## Methods

### Study participants and sampling technique

We recruited 5207 school children between the age of 10 to 20 years, who were attending public or private schools in Sri Lanka, during September 2022 to 31st March 2023 as previously described according to the WHO UNITY protocol [14, 15]. Briefly, children were recruited following informed written consent from the parents/guardians and assent was taken from children. The study was carried out in nine districts in Sri Lanka, representative of each of the nine provinces. A stratified multi-stage cluster sampling method was used to select the schools in each district, with a cluster size of 40 students from each cluster. A probability proportionate to the size (PPS) sampling technique was used to select the sample size from each district, as the population size and urbanicity grade varied in different districts.

A pre-tested and structured interviewer-administered questionnaire was used to record basic demographic details and details of the vaccination history. Blood samples (5ml) were collected by venipuncture by a trained nurse or a medical officer. The samples were centrifuged, serum separated and stored in −80 C freezers, until further use for the assays.

### Ethics statement

The study was approved by the Ethics review Committee of the University of Sri Jayewardenepura, Sri Lanka and also received administrative clearance of the Ministry of Health, Sri Lanka. All subjects and their parents/guardians gave informed written consent.

### Assays for SARS-CoV-2 specific total antibodies and ACE2 blocking antibodies

SARS-COV-2 specific total antibody (IgM, IgG and IgA) responses were evaluated using the Wantai SARS-CoV-2 Ab ELISA (Beijing Wantai Biological Pharmacy Enterprise, China) as previously described. This assay has been used in carrying out serosurveys for SARS-CoV-2 previously in Sri Lanka, in the Colombo district [16]. This assay was shown to have a sensitivity of 98% [17] and was found to be 100% specific in serum samples obtained in 2018, in Sri Lankan individuals. A cut-off value for each ELISA was calculated according to manufacturer’s instructions. Based on the cut-off value, the antibody index was calculated by dividing the absorbance of each sample by the cut-off value, according to the manufacturer’s instructions.

The ACE2 blocking antibodies were measured by using the surrogate Nab test (sVNT) that been widely used as a surrogate measure for the presence of neutralizing antibodies (Nabs) and has widely been used in many studies, including previous studies in Sri Lanka [18–20]. This sVNT assay [18], measures the percentage of inhibition of binding of the RBD of the S protein to recombinant ACE2 (Genscript Biotech, USA). An inhibition percentage ≥ 25% in a sample was considered as positive for ACE2 blocking antibodies. This assay was found to be 100% specific for measuring ACE2 blocking antibodies in the Sri Lankan population [21]. The sVNT assay was only conducted for samples that gave a positive result with the Wantai SARS-CoV-2 Ab ELISA.

### Haemagglutination test (HAT) to detect antibodies to the receptor binding domain (RBD) of omicron variants

The HAT was carried out as previously described using the BA.2.75 and XBB.1.5 versions of the IH4-RBD reagents with additional mutations in the RBD (Y365F, T392W and V395I) [22, 23]. The assays were carried out and interpreted as previously described by us and these assays have been used previously in serosurveys carried out in the Colombo district in Sri Lanka [16, 24]. The HAT assays were performed with sera at 1:40 and 1:80 dilutions, to determine presence of RBD-specific antibodies. The RBD-specific antibody titre for the serum sample was defined by the last well in which the complete absence of “teardrop” formation was observed. A titre of 1:40 was considered as a positive response, as previously described [25]. A HAT titre of 1: 40 was shown to detect 99% of samples which had neutralizing antibody titres of ≥ 20 (50% inhibitory concentrations, IC_50_) assessed with the microneutralization assay [25]. The plates were incubated at room temperature for one hour to allow red blood cells to settle and were then read by tilting the plate for 30Lseconds.

### Assessment of the nutrition status

Anthropometric measurements were obtained at the time the data was collected and blood samples obtained at the schools of the children. The height was measured by a stadiometer to within 0.5cm and weight was measured using a digital scale, which was calibrated regularly throughout the study. In calculating the body mass index centile (BMI) in children aged 10 to 18, the BMI was plotted on the WHO BMI for age growth charts for boys or girls to acquire the percentile ranking, as percentile rankings are the most suitable indicator for growth patterns in children [26]. The BMI centile for age was used instead of Z-score for BMI for age, as it was shown to overestimate the proportion of children with malnutrition in some populations [27].

### Statistical analysis

GraphPad Prism version 10.1 was used for statistical analysis. As the data were not normally distributed, differences in means were compared using the Mann-Whitney U test (two tailed), and the Kruskal-Wallis test was used to compare the differences of the antibody levels between vaccinated and unvaccinated children in the different districts. The Wilcoxon matched-pairs signed rank test was used when comparing paired data, where responses to different omicron variants were compared in the same individuals. Degree of associations between vaccination status and antibodies to SARS-CoV-2 omicron variants, was expressed as the odds ratio (OR), which was obtained from standard contingency table analysis by Haldane’s modification of Woolf’s method. Chi Square tests or the Fisher’s exact test was used to determine the p value.

## Results

### Vaccination uptake rates in children in different districts in Sri Lanka

3119/5207 (59.90%) of children had received at least one dose of the COVID-19, Pfizer BioNTech (BNT162b2) vaccine and 1967/5207 (37.78%) had received two doses. Of those who were eligible to take the vaccine (children ≥ 12 years of age), the overall vaccination rates were 3086/4155 (74.27%). None of the children had received any booster doses, as these were not made available to children under the age of 19 years. Vaccination rates were lowest in the Matara district (47.23%), while the highest vaccination rate was in Polonnaruwa district (68.09%).

The number of children in each age group in each district, who were vaccinated and unvaccinated are shown in supplementary tables 1 to 9.

### Seropositivity rates in vaccinated and unvaccinated children the nine districts in relation to urbanicity

3111/3119 (99.7%) children who had received at least one dose of the vaccine were seropositive for SARS-CoV-2 and 2008/2088 (96.2%) of unvaccinated children (Table 1). Although there was no difference in the seropositivity between these two groups, the vaccinated children had significantly higher (p<0.0001) titres of total antibodies (median 13.5, IQR 12.9 to 14.18 units) compared to unvaccinated children (median 13.3, IQR 12.5 to 13.9 unit). While > 99.4% of vaccinated children were seropositive in all districts, >94.25% of unvaccinated children were also seropositive in all districts. Therefore, overall, in Sri Lanka, irrespective of vaccination status, >94.25% of children were seropositive for SARS-CoV-2, with seropositivity rates reaching > 97% in four districts. There was no difference in the seropositivity rates in unvaccinated children in urban (97.5%), rural (95.8%) and estate (96.2%), showing that children in all areas in Sri Lanka were equally infected with the SARS-CoV-2 virus.

**Table 1:** The number of children recruited from each district from Sri Lanka, with seropositivity rates in vaccinated and unvaccinated children measured by the Wantai total antibody assay.

| **District** | **Number** | **Number who**<br>**received at**<br>**least one dose**<br>**of the vaccine**<br>**N (%)** | **Number who**<br>**received two**<br>**doses of the**<br>**vaccine**<br>**N (%)** | **Number of**<br>**seropositive**<br>**children who**<br>**received at least**<br>**one dose of the**<br>**vaccine**<br>**N (%)** | **Number of**<br>**seropositive**<br>**unvaccinated**<br>**children**<br>**N (%)** |
| --- | --- | --- | --- | --- | --- |
| Trincomalee | 265 | 152 (57.36%) | 109 (41.13%) | 152 (100.00%) | 112 (99.12%) |
| Jaffna | 321 | 213 (66.36%) | 194 (60.44%) | 213 (100.00%) | 105 (97.22%) |
| Kurunegala | 827 | 514 (62.15%) | 255 (30.83%) | 511 (99.42%) | 295 (94.25%) |
| Matara | 506 | 239 (47.23%) | 77 (15.22%) | 238 (99.58%) | 254 (95.13%) |
| Ratnapura | 495 | 316 (63.84%) | 217 (43.84%) | 316 (100.00%) | 176 (98.32%) |
| Polonnaruwa | 257 | 175 (68.09%) | 171 (66.54%) | 175 (100.00%) | 79 (96.34%) |
| Gampaha | 1354 | 732 (54.06%) | 462 (34.12%) | 731 (99.86%) | 595 (95.66%) |
| Kandy | 681 | 456 (66.96%) | 327 (48.02%) | 455 (99.78%) | 220 (97.78%) |
| Badulla | 501 | 322 (64.27%) | 155 (30.94%) | 320 (99.38%) | 172 (96.09%) |
| **Islandwide** | **5207** | **3119 (59.90%)** | **1967 (37.78%)** | **3111 (99.75%)** | **2008 (96.17%)** |

The number of children recruited from each district and seropositivity rates in vaccinated and unvaccinated children are shown in supplementary tables 1 to 9.

### Differences in ACE2 blocking antibody titres in vaccinated and unvaccinated children

2984/3111 (95.9%) children who had received at least one dose of the vaccine had ACE2 blocking antibodies above the cut-off threshold of a positive response (>25% inhibition), compared to 1346/2008 (67.0%) of unvaccinated children. The positivity rates for ACE2 blocking antibodies of vaccinated children from all districts was >91.4%, with positivity rates being >96% in five districts (Table 2). Unvaccinated children (median 67.6%, IQR 16.8 to 97.3 % of inhibition) had significantly lower (p<0.0001) titres than vaccinated children (median 98.7, IQR 91.9 to 99.2 % of inhibition). Interestingly, the ACE2 blocking antibody positivity rates and antibody titres were higher in some districts compared to others, among unvaccinated children (Table 2, Figure 1A and B). For instance, unvaccinated children in Jaffna had a median ACE2 blocking antibody titres of 91.1 (IQR 48.6 to 98.7% of inhibition). The ACE2 blocking antibody titres in vaccinated children were also highest in the Jaffna district followed by the Trinco district.

**Figure 1:**
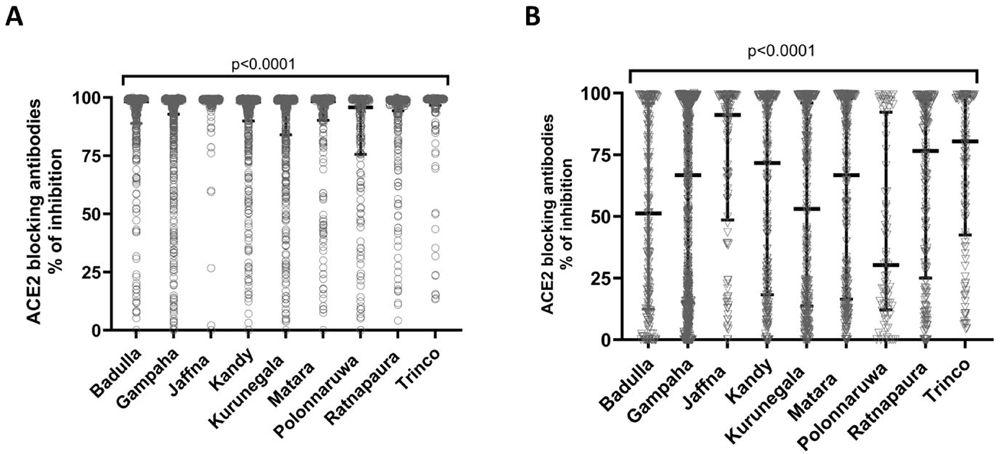
ACE2 blocking antibody levels determined by the surrogate SARS-CoV-2 neutralising antibody assay (sVNT) in vaccinated and unvaccinated children of different districts in Sri Lanka. The ACE2 blocking antibody levels (% of inhibition) were measured in vaccinated (A, n=3111) and unvaccinated (B, n=2008) children between the ages of 10-20 years of age in nine districts in Sri Lanka. The Mann-Whitney U test (two tailed) was used to calculate the differences in the means in the ACE2 antibody titres in different districts in vaccinated and unvaccinated children. All tests were two sided. Data are presented as median values +/- interquartile ranges as appropriate.

**Table 2:** The number of children tested for ACE2 blocking antibodies from each district in Sri Lanka, with seropositivity rates in vaccinated and unvaccinated children.

| **District** | **Total Number of seropositive children tested in each district** | **Number of children who had detectable ACE2 blocking antibodies who received at least one dose of the vaccine N (%)** | **Number of unvaccinated children who had detectable ACE2 blocking antibodies N (%)** |
| --- | --- | --- | --- |
| Trincomalee | 264 | 146 (96.05%) | 92 (82.14%) |
| Jaffna | 318 | 210 (98.59%) | 85 (80.95%) |
| Kurunegala | 806 | 490 (95.89%) | 178 (60.34%) |
| Matara | 492 | 225 (94.54%) | 166 (65.35%) |
| Ratnapura | 492 | 308 (97.47%) | 132 (75.00%) |
| Polonnaruwa | 254 | 160 (91.43%) | 42 (53.16%) |
| Gampaha | 1326 | 704 (96.31%) | 387 (65.04%) |
| Kandy | 675 | 439 (96.48%) | 153 (69.55%) |
| Badulla | 492 | 302 (94.38%) | 111 (64.53%) |
| **Islandwide** | **5119** | **2984 (95.92%)** | **1346 (67.03%)** |

The positivity rates for ACE2 blocking antibodies were significantly higher in unvaccinated children in urban (291/391, 74.4%, p=0.0016) and estate areas (37/48, 77.1%, p=0.004), compared to children living in rural areas (1018/1569, 64.9%). However, there was no association with the population density and ACE2 blocking antibody levels in each district (Spearmans’ R=0.15, p=0.69). The positivity rates for ACE2 blocking antibodies in vaccinated and unvaccinated children are shown in supplementary tables 10 to 18.

### Antibody responses to SARS-CoV-2 omicron variants in vaccinated and unvaccinated children

We carried out HAT assays to assess antibody responses to BA.2.75 and XBB.1.5 in samples of 10% of the unvaccinated seropositive children, which were randomly selected representative of all the nine districts (n=202). These assays were carried out in a similar number (n=202) of vaccinated children representative of all nine districts in Sri Lanka. 130/202 (64.3%) vaccinated and 55 (27.2 %) unvaccinated children had an antibody titre of ≥ 1:40 to BA.2.75. For XBB.1.5, 87 (43.1%) vaccinated and 62 (30.7%) unvaccinated children had an antibody titre of ≥ 1:40. The vaccinated children had significantly higher antibody titres to both XBB.1.5 (p=0.01) and BA.2.75 (p<0.0001), although this difference was more significant for BA.2.75 (Figure 2). In vaccinees, the antibody titres were significantly higher to BA.2.75 than to XBB.1.5 (p<0.0001) whereas there was no difference in antibody titres to these variants in unvaccinated children (p=0.18) (Figure 2). Therefore, the vaccinated children were significantly more likely to have a positive response of ≥ 1:40 to BA.2.75 (odds ratio 2.4, 95% CI 1.6 to 3.6, p<0.0001) compared to XBB.1.5. In fact, 59 (29.2%) of vaccinated children had a titre of ≥ 1:80 to BA.2.75 compared to 35 (17.3%) to XBB.1.5.

**Figure 2:**
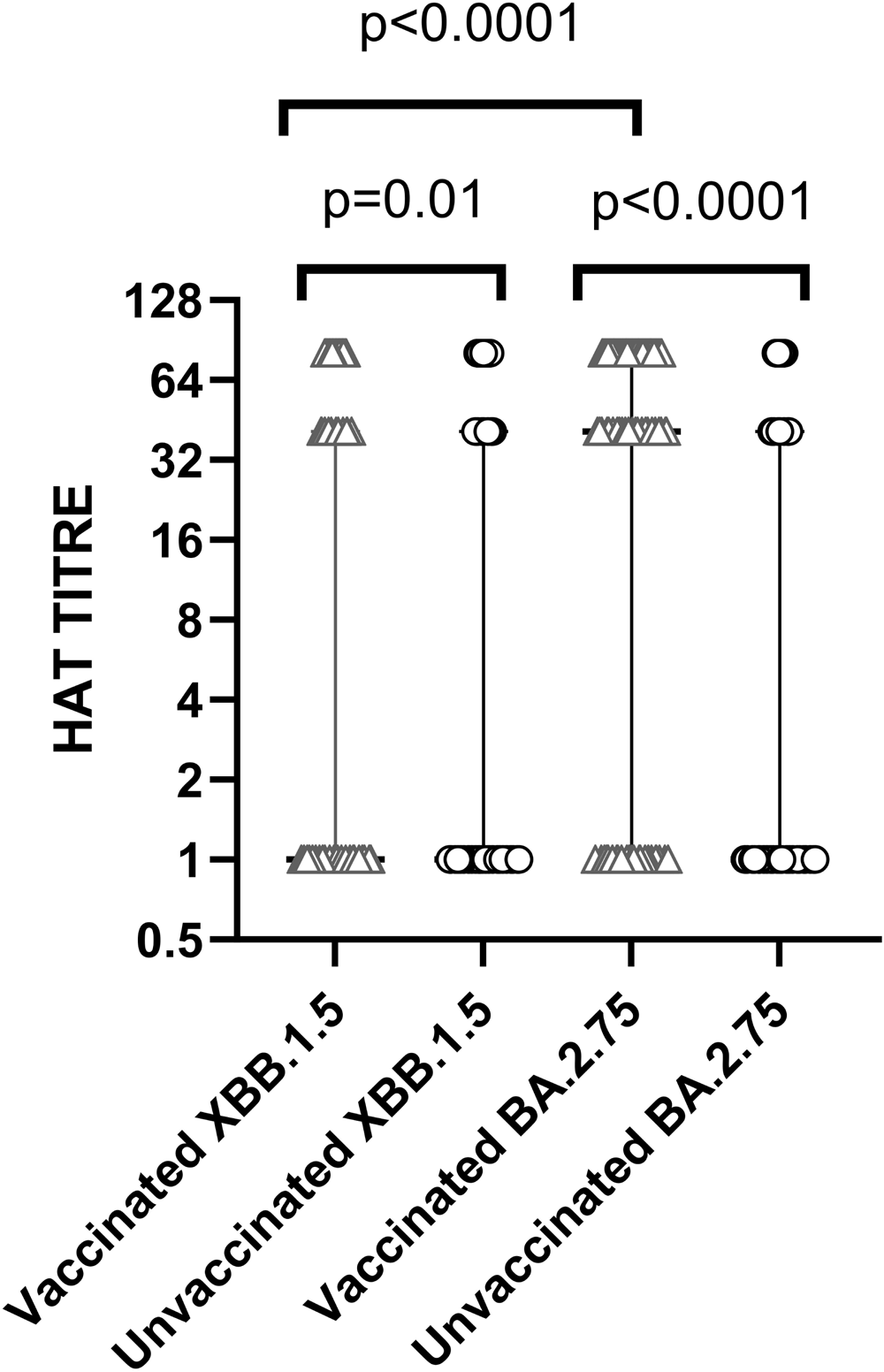
Antibody responses to the omicron sub-lineages XBB.1.5 and BA.2.75 in vaccinated and unvaccinated children in Sri Lanka. The hemagglutination test was carried out to determine antibody responses to XBB.1.5 and BA.2.75 in vaccinated (n=202) and unvaccinated (n=202) representative of all nine districts of Sri Lanka, at 1:40 and 1:80 dilutions, as previously described. The Wilcoxon matched pairs signed rank test was used to compare the means of the HAT titres for the XBB.1.5 and BA.2.75 in the same individuals. The Mann-Whitney U test (two tailed) was used to calculate the differences in the means in the HAT titres in the vaccinated and unvaccinated children. All tests were two sided. Data are presented as median values +/- interquartile ranges as appropriate.

### The magnitude of antibody responses to SARS-CoV-2 based on nutritional status

As previously described by us [28], in this island wide large cohort of children, 4782/5207, were children between the ages of 10 to 18, and the BMI centile was determined to find out their nutritional status. In this cohort of children (n=4782), 1057 (22.1%) had a BMIs <3rd centile for age, and therefore were classified as underweight. 215 (4.5%) children had a BMI of >97th centile for age, when plotted on the WHO BMI for age charts and were considered severely overweight. Among the unvaccinated children, those who had a BMI of <3^rd^ centile had significantly lower ACE blocking antibodies (median 51.9, IQR 13.1 to 95.5 % of inhibition), which is a surrogate marker for the presence of Nabs compared to children of other categories (Figure 3A). Children with a BMI centile between 85^th^ to 97^th^ had the higher titres of ACE2 blocking antibodies (median 82.7, IQR 21.6 to 99.7, % of inhibition). Among vaccinated children the ACE2 blocking antibody titres were similar in children of different nutritional status (Figure 3B).

**Figure 3:**
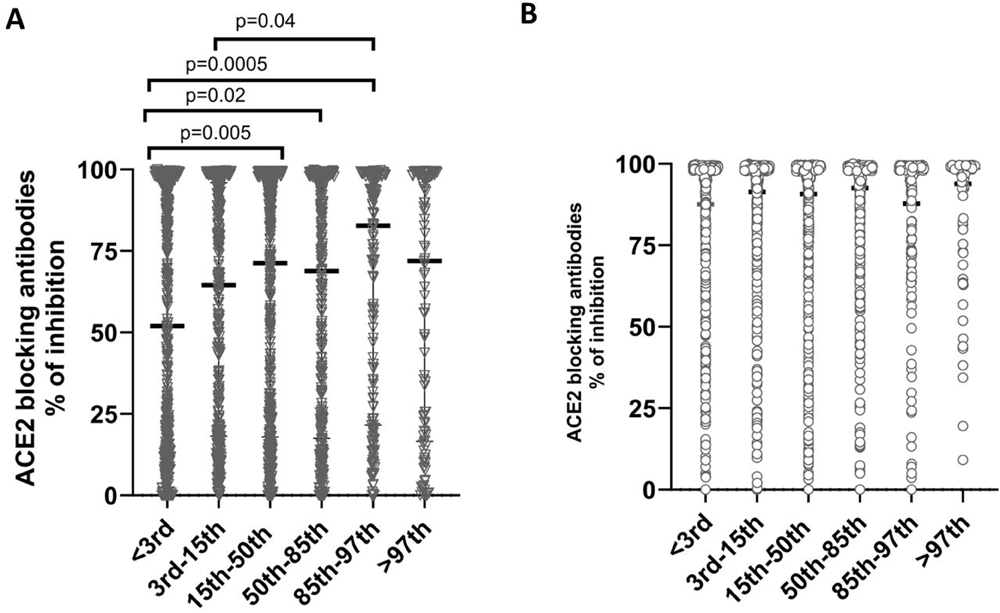
ACE2 blocking antibody levels determined by the surrogate SARS-CoV-2 neutralising antibody assay (sVNT) in vaccinated and unvaccinated children of different BMI centile categories. The ACE2 blocking antibody levels (% of inhibition) were measured in vaccinated (A, n=2723) and unvaccinated (B, n=1973) children between the ages of 10-18 years of age, of different BMI centiles for age. The Mann-Whitney U test (two tailed) was used to calculate the differences in the means in the ACE2 antibody titres in different BMI groups in the vaccinated and unvaccinated. All tests were two sided. Data are presented as median values +/- interquartile ranges as appropriate.

## Discussion

In this study we carried out an island-wide serosurvey to determine the SARS-CoV-2 seropositivity rates among vaccinated and unvaccinated children, representing all the nine provinces in Sri Lanka. We found that the overall seropositivity rates of the unvaccinated children (96.2%) and vaccinated children (99.7%) were similar, indicating a high infection rate in all areas in Sri Lanka by March 2023. However, the titres of ACE2 blocking antibodies (surrogate marker for Nabs) and the levels of total antibodies to the RBD of the virus was significantly higher among vaccinated children, indicating vaccines provided a higher magnitude of antibodies compared to natural infection.

Interestingly, the ACE2 antibody positivity rates and titres were significantly higher among unvaccinated children in some districts compared to others. We previously showed that in unvaccinated, naturally infected adults, many had low levels of ACE2 blocking antibodies (median levels, 22.3 % of inhibition), with only 50% having antibody levels above the positive cut-off threshold (>25% of inhibition) [21]. In this study, overall, 67.0% of unvaccinated children had ACE2 blocking antibody titres above the cut-off threshold and the median values were 67.6%, which were several folds higher than what we found in individuals who had one natural infection with SARS-Cov-2 in 2020 [21]. In some districts such as Jaffna the median values were 91.1, followed by 80.4 % of inhibition in the Trinco district, which are several folds higher than found in individuals following a single episode of natural infection. Therefore, given the high positivity rates for ACE2 blocking antibodies and high titres seen in unvaccinated children, it is likely that children (vaccinated and unvaccinated) are likely to have been infected more than once with SARS-CoV-2.

We assessed antibody responses to omicron sub-lineages BA.2.75 and XBB.1.5 in the sub cohort of vaccinated and unvaccinated children. 64.3% of vaccinated and 27.2 % unvaccinated children had antibody titre above the positive cut off threshold to BA.2.75, and 43.1% of vaccinated and 30.7% unvaccinated children to XBB.1.5. Sri Lanka reported circulation of BA.2.75 variants during the latter part of 2022 [2], while the XBB variants were only found after the study recruitment had finished. Therefore, although the children could have experienced infection with BA.2.75, they are less likely to have been exposed to the variants of the XBB lineage. However, there was no difference in positivity rates or the antibody titres to BA.2.75 and XBB.1.5 in unvaccinated children, suggesting that previous infection in many induced a broader antibody response to other variants. Vaccinated children had higher positivity rates and higher antibody titres to both BA.2.75 and XBB.1.5 than unvaccinated children, showing that vaccination induced a broader antibody response of higher magnitude for many SARS-CoV-2 variants. Interestingly, the vaccinated children had significantly higher antibody responses to BA.2.75 than to XBB.1.5. As XBB.1.5 has many more mutations within the RBD than BA.2.75 [9], it is likely that it escapes vaccine induce immunity at a far greater extent than BA.2.75. Furthermore, many children received their vaccines during the BA.2 wave in Sri Lanka [2], it is likely they would have been exposed to the vaccine virus and BA.2 during a very short period, thereby inducing robust immune responses to BA.2 sub-lineages. Indeed, Sri Lanka has reported very few cases of COVID-19 in 2023, possibly due to limited testing carried out [8]. However, the number have deaths have also remained low, with 69 deaths reported for 2023, until 20^th^ November 2023 [8]. Therefore, vaccination along with natural infection with omicron variants such as BA.2 and subsequently BA.4/BA.5, would have given the population a broader immune response, thereby reducing the impact of the newer omicron sub lineages of the XBB series, despite the non-availability of bi-valent or monovalent vaccines with omicron boosters.

Sri Lanka is going through an economic crisis, with many children not having access to nutritious food [13]. Indeed, we found that 22.1% of children were <3^rd^ BMI centile for age, indicating under nutrition. Although the seropositivity rates among different age groups did not differ, there were significant differences in ACE2 blocking antibody levels, which are surrogate markers of Nabs [18, 21]. Nabs antibodies prevent binding to the ACE2 receptor and have shown to associate with protection [29]. Children with undernutrition (<3^rd^ BMI centile for age), had significantly lower ACE2 blocking antibody titres compared to children of healthy weight. There was no difference in the ACE2 blocking antibodies in children with undernutrition and obese children (>9^th^ BMI centile for age), suggesting that those with a normal nutrition status, are likely to have a more robust antibody response. However, in this study we only investigated antibody responses to the spike protein using different types of assays and it would be important to understand the antibody responses to other proteins such as the N protein, which has also shown to associate with protection. Furthermore, although more recent SARS-CoV-2 omicron variants almost completely evade neutralization with antibodies specific to the earlier SARS-CoV-2 variants (Wuhan-Hu-1) [9], they do not completely evade T cell responses [30, 31]. Therefore, in order to fully understand the population immunity to SARS-CoV-2 variants, also in the context of the nutrition status, it would be important to assess the functionality, magnitude and the breadth of T and B cell responses to the virus.

## Funding statement

This study has been supported by WHO Unity Studies, a global sero-epidemiological standardization initiative, with funding to WHO and the UK Medical Research Council. T.K.T. is funded by the Townsend-Jeantet Charitable Trust (charity number 1011770) and the EPA Cephalosporin Early Career Researcher Fund. A.T. are funded by the Chinese Academy of Medical Sciences (CAMS) Innovation Fund for Medical Science (CIFMS), China (grant no. 2018-I2M-2-002).

## Data Availability

All data are available within the manuscript, figures and the tables. Individual data points are shown in all figures.

## Notes

### Competing Interest Statement

The authors have declared no competing interest.

