## Supplementary tables for "The magnitude and cross reactivity of SARS-CoV-2 specific antibody responses in vaccinated and unvaccinated Sri Lankan children and association with nutrition status"

**Supplementary Data**

| **Age Group** | **Total Number in Each Age Group** | **Seropositivity Rate Based on Detection of Total Antibodies N (%)** | **Number Vaccinated N (%)** | **Number Vaccinated Seropositive N (%)** | **Number Unvaccinated Seropositive N (%)** |
| --- | --- | --- | --- | --- | --- |
| **10** | 25 | 25 (100.00%) | 0 (0.00%) | 0 | 25 (100.00%) |
| **11** | 28 | 27 (96.43%) | 1 (3.57%) | 1 (100.00%) | 26 (96.30%) |
| **12** | 29 | 29 (100.00%) | 4 (13.79%) | 4 (100.00%) | 25 (100.00%) |
| **13** | 31 | 31 (100.00%) | 23 (74.19%) | 23 (100.00%) | 8 (100.00%) |
| **14** | 29 | 29 (100.00%) | 21 (72.41%) | 21 (100.00%) | 8 (100.00%) |
| **15** | 39 | 39 (100.00%) | 32 (82.05%) | 32 (100.00%) | 7 (100.00%) |
| **16** | 26 | 26 (100.00%) | 19 (73.08%) | 19 (100.00%) | 7 (100.00%) |
| **17-18** | 29 | 29 (100.00%) | 26 (89.66%) | 26 (100.00%) | 3 (100.00%) |
| **19-20** | 29 | 29 (100.00%) | 26 (89.66%) | 26 (100.00%) | 3 (100.00%) |
| **Total** | **265** | **264 (99.62%)** | **152 (57.36%)** | **152 (100.00%)** | **112 (99.12%)** |

**Table 1: The number of children in each age group, seropositivity rates based on total antibodies and vaccination rates for SARS-CoV-2 in Trincomalee**

| **Age Group** | **Total Number in Each Age Group** | **Seropositivity Rate Based on Detection of Total Antibodies N (%)** | **Number Vaccinated N (%)** | **Number Vaccinated Seropositive N (%)** | **Number Unvaccinated Seropositive N (%)** |
| --- | --- | --- | --- | --- | --- |
| **10** | 16 | 15 (93.75%) | 0 (0.00%) | 0 | 15 (93.75%) |
| **11** | 19 | 18 (94.74%) | 0 (0.00%) | 0 | 18 (94.74%) |
| **12** | 27 | 26 (96.30%) | 1 (3.70%) | 1 (100.00%) | 25 (96.15%) |
| **13** | 32 | 32 (100.00%) | 28 (87.50%) | 28 (100.00%) | 4 (100.00%) |
| **14** | 27 | 27 (100.00%) | 21 (77.78%) | 21 (100.00%) | 6 (100.00%) |
| **15** | 28 | 28 (100.00%) | 22 (78.57%) | 22 (100.00%) | 6 (100.00%) |
| **16** | 28 | 28 (100.00%) | 26 (92.86%) | 26 (100.00%) | 2 (100.00%) |
| **17-18** | 54 | 54 (100.00%) | 51 (94.44%) | 51 (100.00%) | 3 (100.00%) |
| **19-20** | 26 | 26 (100.00%) | 26 (100.00%) | 26 (100.00%) | 0 |
| **Total** | **257** | **254 (98.83%)** | **175 (68.09%)** | **175 (100.00%)** | **79 (96.34%)** |

**Table 2: The number of children in each age group, seropositivity rates based on total antibodies and vaccination rates for SARS-CoV-2 in Polonnaruwa**

| **Age Group** | **Total Number in Each Age Group** | **Seropositivity Rate Based on Detection of Total Antibodies N (%)** | **Number Vaccinated N (%)** | **Number Vaccinated Seropositive N (%)** | **Number Unvaccinated Seropositive N (%)** |
| --- | --- | --- | --- | --- | --- |
| **10** | 56 | 52 (92.86%) | 1 (1.79%) | 1 (100.00%) | 51 (92.73%) |
| **11** | 53 | 48 (90.57%) | 1 (1.89%) | 0 (0.00%) | 48 (92.31%) |
| **12** | 48 | 45 (93.75%) | 5 (10.42%) | 5 (100.00%) | 40 (93.02%) |
| **13** | 58 | 58 (100.00%) | 37 (63.79%) | 37 (100.00%) | 21 (100.00%) |
| **14** | 52 | 51 (98.08%) | 32 (61.54%) | 32 (100.00%) | 19 (95.00%) |
| **15** | 58 | 57 (98.28%) | 31 (53.45%) | 31 (100.00%) | 26 (96.30%) |
| **16** | 55 | 55 (100.00%) | 31 (56.36%) | 31 (100.00%) | 24 (100.00%) |
| **17-18** | 56 | 56 (100.00%) | 44 (78.57%) | 44 (100.00%) | 12 (100.00%) |
| **19-20** | 70 | 70 (100.00%) | 57 (81.43%) | 57 (100.00%) | 13 (100.00%) |
| **Total** | **506** | **492 (97.23%)** | **239 (47.23%)** | **238 (99.58%)** | **254 (95.13%)** |

**Table 3: The number of children in each age group, seropositivity rates based on total antibodies and vaccination rates for SARS-CoV-2 in Matara**

| **Age Group** | **Total Number in Each Age Group** | **Seropositivity Rate Based on Detection of Total Antibodies N (%)** | **Number Vaccinated N (%)** | **Number Vaccinated Seropositive N (%)** | **Number Unvaccinated Seropositive N (%)** |
| --- | --- | --- | --- | --- | --- |
| **10** | 56 | 54 (96.43%) | 2 (3.57%) | 2 (100.00%) | 52 (96.30%) |
| **11** | 54 | 54 (100.00%) | 1 (1.85%) | 1 (100.00%) | 53 (100.00%) |
| **12** | 46 | 45 (97.83%) | 6 (13.04%) | 6 (100.00%) | 39 (97.50%) |
| **13** | 57 | 57 (100.00%) | 45 (78.95%) | 45 (100.00%) | 12 (100.00%) |
| **14** | 54 | 54 (100.00%) | 50 (92.59%) | 50 (100.00%) | 4 (100.00%) |
| **15** | 55 | 55 (100.00%) | 51 (92.73%) | 51 (100.00%) | 4 (100.00%) |
| **16** | 57 | 57 (100.00%) | 49 (85.96%) | 49 (100.00%) | 8 (100.00%) |
| **17-18** | 86 | 86 (100.00%) | 84 (97.67%) | 84 (100.00%) | 2 (100.00%) |
| **19-20** | 30 | 30 (100.00%) | 28 (93.33%) | 28 (100.00%) | 2 (100.00%) |
| **Total** | **495** | **492 (99.39%)** | **316 (63.84%)** | **316 (100.00%)** | **176 (98.32%)** |

**Table 4: The number of children in each age group, seropositivity rates based on total antibodies and vaccination rates for SARS-CoV-2 in Ratnapura**

| **Age Group** | **Total Number in Each Age Group** | **Seropositivity Rate Based on Detection of Total Antibodies N (%)** | **Number Vaccinated N (%)** | **Number Vaccinated Seropositive N (%)** | **Number Unvaccinated Seropositive N (%)** |
| --- | --- | --- | --- | --- | --- |
| **10** | 28 | 28 (100.00%) | 0 (0.00%) | 0 | 28 (100.00%) |
| **11** | 36 | 35 (97.22%) | 2 (5.56%) | 2 (100.00%) | 33 (97.06%) |
| **12** | 30 | 29 (96.67%) | 5 (16.67%) | 5 (100.00%) | 24 (96.00%) |
| **13** | 35 | 35 (100.00%) | 30 (85.71%) | 30 (100.00%) | 5 (100.00%) |
| **14** | 34 | 34 (100.00%) | 27 (79.41%) | 27 (100.00%) | 7 (100.00%) |
| **15** | 28 | 27 (96.43%) | 25 (89.29%) | 25 (100.00%) | 2 (66.67%) |
| **16** | 45 | 45 (100.00%) | 42 (93.33%) | 42 (100.00%) | 3 (100.00%) |
| **17-18** | 61 | 61 (100.00%) | 59 (96.72%) | 59 (100.00%) | 2 (100.00%) |
| **19-20** | 24 | 24 (100.00%) | 23 (95.83%) | 23 (100.00%) | 1 (100.00%) |
| **Total** | **321** | **318 (99.07%)** | **213 (66.36%)** | **213 (100.00%)** | **105 (97.22%)** |

**Table 5: The number of children in each age group, seropositivity rates based on total antibodies and vaccination rates for SARS-CoV-2 in Jaffna**

| **Age Group** | **Total Number in Each Age Group** | **Seropositivity Rate Based on Detection of Total Antibodies N (%)** | **Number Vaccinated N (%)** | **Number Vaccinated Seropositive N (%)** | **Number Unvaccinated Seropositive N (%)** |
| --- | --- | --- | --- | --- | --- |
| **10** | 87 | 83 (95.40%) | 3 (3.45%) | 3 (100.00%) | 80 (95.24%) |
| **11** | 90 | 83 (92.22%) | 11 (12.22%) | 11 (100.00%) | 72 (91.14%) |
| **12** | 85 | 80 (94.12%) | 14 (16.47%) | 14 (100.00%) | 66 (92.96%) |
| **13** | 96 | 95 (98.96%) | 77 (80.21%) | 77 (100.00%) | 18 (94.74%) |
| **14** | 93 | 92 (98.92%) | 78 (83.87%) | 77 (98.72%) | 15 (100.00%) |
| **15** | 92 | 91 (98.91%) | 70 (76.09%) | 70 (100.00%) | 21 (95.45%) |
| **16** | 84 | 83 (98.81%) | 72 (85.71%) | 71 (98.61%) | 12 (100.00%) |
| **17-18** | 130 | 129 (99.23%) | 121 (93.08%) | 120 (99.17%) | 9 (100.00%) |
| **19-20** | 70 | 70 (100.00%) | 68 (97.14%) | 68 (100.00%) | 2 (100.00%) |
| **Total** | **827** | **806 (97.46%)** | **514 (62.15%)** | **511 (99.42%)** | **295 (94.25%)** |

**Table 6: The number of children in each age group, seropositivity rates based on total antibodies and vaccination rates for SARS-CoV-2 in Kurunegala**

| **Age Group** | **Total Number in Each Age Group** | **Seropositivity Rate Based on Detection of Total Antibodies N (%)** | **Number Vaccinated N (%)** | **Number Vaccinated Seropositive N (%)** | **Number Unvaccinated Seropositive N (%)** |
| --- | --- | --- | --- | --- | --- |
| **10** | 36 | 34 (94.44%) | 2 (5.56%) | 1 (50.00%) | 33 (97.06%) |
| **11** | 80 | 79 (98.75%) | 3 (3.75%) | 3 (100.00%) | 76 (98.70%) |
| **12** | 69 | 66 (95.65%) | 14 (20.29%) | 14 (100.00%) | 52 (94.55%) |
| **13** | 74 | 74 (100.00%) | 60 (81.08%) | 60 (100.00%) | 14 (100.00%) |
| **14** | 76 | 76 (100.00%) | 66 (86.84%) | 66 (100.00%) | 10 (100.00%) |
| **15** | 84 | 84 (100.00%) | 71 (84.52%) | 71 (100.00%) | 13 (100.00%) |
| **16** | 100 | 100 (100.00%) | 91 (91.00%) | 91 (100.00%) | 9 (100.00%) |
| **17-18** | 89 | 89 (100.00%) | 85 (95.51%) | 85 (100.00%) | 4 (100.00%) |
| **19-20** | 73 | 73 (100.00%) | 64 (87.67%) | 64 (100.00%) | 9 (100.00%) |
| **Total** | **681** | **675 (99.12%)** | **456 (66.96%)** | **455 (99.78%)** | **220 (97.78%)** |

**Table 7: The number of children in each age group, seropositivity rates based on total antibodies and vaccination rates for SARS-CoV-2 in Kandy**

| **Age Group** | **Total Number in Each Age Group** | **Seropositivity Rate Based on Detection of Total Antibodies N (%)** | **Number Vaccinated N (%)** | **Number Vaccinated Seropositive N (%)** | **Number Unvaccinated Seropositive N (%)** |
| --- | --- | --- | --- | --- | --- |
| **10** | 130 | 120 (92.31%) | 1 (0.77%) | 1 (100.00%) | 119 (92.25%) |
| **11** | 144 | 138 (95.83%) | 1 (0.69%) | 1 (100.00%) | 137 (95.80%) |
| **12** | 150 | 144 (96.00%) | 14 (9.33%) | 14 (100.00%) | 130 (95.59%) |
| **13** | 156 | 155 (99.36%) | 101 (64.74%) | 101 (100.00%) | 54 (98.18%) |
| **14** | 149 | 148 (99.33%) | 104 (69.80%) | 104 (100.00%) | 44 (97.78%) |
| **15** | 157 | 156 (99.36%) | 110 (70.06%) | 110 (100.00%) | 46 (97.87%) |
| **16** | 144 | 143 (99.31%) | 97 (67.36%) | 97 (100.00%) | 46 (97.87%) |
| **17-18** | 244 | 243 (99.59%) | 227 (93.03%) | 227 (100.00%) | 16 (94.12%) |
| **19-20** | 80 | 79 (98.75%) | 77 (96.25%) | 76 (98.70%) | 3 (100.00%) |
| **Total** | **1354** | **1326 (97.93%)** | **732 (54.06%)** | **731 (99.86%)** | **595 (95.66%)** |

**Table 8: The number of children in each age group, seropositivity rates based on total antibodies and vaccination rates for SARS-CoV-2 in Gampaha**

| **Age Group** | **Total Number in Each Age Group** | **Seropositivity Rate Based on Detection of Total Antibodies N (%)** | **Number Vaccinated N (%)** | **Number Vaccinated Seropositive N (%)** | **Number Unvaccinated Seropositive N (%)** |
| --- | --- | --- | --- | --- | --- |
| **10** | 60 | 58 (96.67%) | 4 (6.67%) | 4 (100.00%) | 54 (96.43%) |
| **11** | 54 | 53 (98.15%) | 0 (0.00%) | 0 | 53 (98.15%) |
| **12** | 60 | 56 (93.33%) | 11 (18.33%) | 11 (100.00%) | 45 (91.84%) |
| **13** | 45 | 45 (100.00%) | 42 (93.33%) | 42 (100.00%) | 3 (100.00%) |
| **14** | 63 | 63 (100.00%) | 61 (96.83%) | 61 (100.00%) | 2 (100.00%) |
| **15** | 64 | 64 (100.00%) | 56 (87.50%) | 56 (100.00%) | 8 (100.00%) |
| **16** | 49 | 49 (100.00%) | 49 (100.00%) | 49 (100.00%) | 0 |
| **17-18** | 83 | 82 (98.80%) | 78 (93.98%) | 77 (98.72%) | 5 (100.00%) |
| **19-20** | 23 | 22 (95.65%) | 21 (91.30%) | 20 (95.24%) | 2 (100.00%) |
| **Total** | **501** | **492 (98.20%)** | **322 (64.27%)** | **320 (99.38%)** | **172 (96.09%)** |

**Table 9: The number of children in each age group, seropositivity rates based on total antibodies and vaccination rates for SARS-CoV-2 in Badulla**

| **Age Group** | **Total Number in Each Age Group** | **Seropositivity Rate By Prescence of ACE2 Blocking Antibodies N (%)** | **Number Vaccinated N (%)** | **Number Vaccinated Seropositive N (%)** | **Number Unvaccinated Seropositive N (%)** |
| --- | --- | --- | --- | --- | --- |
| **10** | 25 | 19 (76.00%) | 0 (0.00%) | 0 | 19 (76.00%) |
| **11** | 27 | 19 (70.37%) | 1 (3.70%) | 1 (100.00%) | 18 (69.23%) |
| **12** | 29 | 25 (86.21%) | 4 (13.79%) | 4 (100.00%) | 21 (84.00%) |
| **13** | 31 | 30 (96.77%) | 23 (74.19%) | 22 (95.65%) | 8 (100.00%) |
| **14** | 29 | 27 (93.10%) | 21 (72.41%) | 19 (90.48%) | 8 (100.00%) |
| **15** | 39 | 39 (100.00%) | 32 (82.05%) | 32 (100.00%) | 7 (100.00%) |
| **16** | 26 | 24 (92.31%) | 19 (73.08%) | 18 (94.74%) | 6 (85.71%) |
| **17-18** | 29 | 27 (93.10%) | 26 (89.66%) | 25 (96.15%) | 2 (66.67%) |
| **19-20** | 29 | 28 (96.55%) | 26 (89.66%) | 25 (96.15%) | 3 (100.00%) |
| **Total** | **264** | **238 (90.15%)** | **152 (57.58%)** | **146 (96.05%)** | **92 (82.14%)** |

**Table 10: The number of children in each age group, seropositivity rates based on ACE2 blocking antibodies and vaccination rates for SARS-CoV-2 in Trincomalee**

| **Age Group** | **Total Number in Each Age Group** | **Seropositivity Rate By Prescence of ACE2 Blocking Antibodies N (%)** | **Number Vaccinated N (%)** | **Number Vaccinated Seropositive N (%)** | **Number Unvaccinated Seropositive N (%)** |
| --- | --- | --- | --- | --- | --- |
| **10** | 15 | 6 (40.00%) | 0 (0.00%) | 0 | 6 (40.00%) |
| **11** | 18 | 10 (55.56%) | 0 (0.00%) | 0 | 10 (55.56%) |
| **12** | 26 | 15 (57.69%) | 1 (3.85%) | 1 (100.00%) | 14 (56.00%) |
| **13** | 32 | 27 (84.38%) | 28 (87.50%) | 25 (89.29%) | 2 (50.00%) |
| **14** | 27 | 23 (85.19%) | 21 (77.78%) | 19 (90.48%) | 4 (66.67%) |
| **15** | 28 | 25 (89.29%) | 22 (78.57%) | 22 (100.00%) | 3 (50.00%) |
| **16** | 28 | 24 (85.71%) | 26 (92.86%) | 24 (92.31%) | 0 (0.00%) |
| **17-18** | 54 | 48 (88.89%) | 51 (94.44%) | 45 (88.24%) | 3 (100.00%) |
| **19-20** | 26 | 24 (92.31%) | 26 (100.00%) | 24 (92.31%) | 0 |
| **Total** | **254** | **202 (79.53%)** | **175 (68.90%)** | **160 (91.43%)** | **42 (53.16%)** |

**Table 11: The number of children in each age group, seropositivity rates based on ACE2 blocking antibodies and vaccination rates for SARS-CoV-2 in Polonnaruwa**

| **Age Group** | **Total Number in Each Age Group** | **Seropositivity Rate By Prescence of ACE2 Blocking Antibodies N (%)** | **Number Vaccinated N (%)** | **Number Vaccinated Seropositive N (%)** | **Number Unvaccinated Seropositive N (%)** |
| --- | --- | --- | --- | --- | --- |
| **10** | 52 | 28 (53.85%) | 1 (1.92%) | 0 (0.00%) | 28 (54.90%) |
| **11** | 48 | 31 (64.58%) | 0 (0.00%) | 0 | 31 (64.58%) |
| **12** | 45 | 28 (62.22%) | 5 (11.11%) | 5 (100.00%) | 23 (57.50%) |
| **13** | 58 | 50 (86.21%) | 37 (63.79%) | 33 (89.19%) | 17 (80.95%) |
| **14** | 51 | 45 (88.24%) | 32 (62.75%) | 32 (100.00%) | 13 (68.42%) |
| **15** | 57 | 47 (82.46%) | 31 (54.39%) | 29 (93.55%) | 18 (69.23%) |
| **16** | 55 | 45 (81.82%) | 31 (56.36%) | 30 (96.77%) | 15 (62.50%) |
| **17-18** | 56 | 51 (91.07%) | 44 (78.57%) | 41 (93.18%) | 10 (83.33%) |
| **19-20** | 70 | 66 (94.29%) | 57 (81.43%) | 55 (96.49%) | 11 (84.62%) |
| **Total** | **492** | **391 (79.47%)** | **238 (48.37%)** | **225 (94.54%)** | **166 (65.35%)** |

**Table 12: The number of children in each age group, seropositivity rates based on ACE2 blocking antibodies and vaccination rates for SARS-CoV-2 in Matara**

| **Age Group** | **Total Number in Each Age Group** | **Seropositivity Rate By Prescence of ACE2 Blocking Antibodies N (%)** | **Number Vaccinated N (%)** | **Number Vaccinated Seropositive N (%)** | **Number Unvaccinated Seropositive N (%)** |
| --- | --- | --- | --- | --- | --- |
| **10** | 54 | 39 (72.22%) | 2 (3.70%) | 1 (50.00%) | 38 (73.08%) |
| **11** | 54 | 38 (70.37%) | 1 (1.85%) | 1 (100.00%) | 37 (69.81%) |
| **12** | 45 | 38 (84.44%) | 6 (13.33%) | 6 (100.00%) | 32 (82.05%) |
| **13** | 57 | 54 (94.74%) | 45 (78.95%) | 44 (97.78%) | 10 (83.33%) |
| **14** | 54 | 53 (98.15%) | 50 (92.59%) | 50 (100.00%) | 3 (75.00%) |
| **15** | 55 | 52 (94.55%) | 51 (92.73%) | 49 (96.08%) | 3 (75.00%) |
| **16** | 57 | 53 (92.98%) | 49 (85.96%) | 47 (95.92%) | 6 (75.00%) |
| **17-18** | 86 | 85 (98.84%) | 84 (97.67%) | 83 (98.81%) | 2 (100.00%) |
| **19-20** | 30 | 28 (93.33%) | 28 (93.33%) | 27 (96.43%) | 1 (50.00%) |
| **Total** | **492** | **440 (89.43%)** | **316 (64.23%)** | **308 (97.47%)** | **132 (75.00%)** |

**Table 13: The number of children in each age group, seropositivity rates based on ACE2 blocking antibodies and vaccination rates for SARS-CoV-2 in Ratnapura**

| **Age Group** | **Total Number in Each Age Group** | **Seropositivity Rate By Prescence of ACE2 Blocking Antibodies N (%)** | **Number Vaccinated N (%)** | **Number Vaccinated Seropositive N (%)** | **Number Unvaccinated Seropositive N (%)** |
| --- | --- | --- | --- | --- | --- |
| **10** | 28 | 21 (75.00%) | 0 (0.00%) | 0 | 21 (75.00%) |
| **11** | 35 | 29 (82.86%) | 2 (5.71%) | 2 (100.00%) | 27 (81.82%) |
| **12** | 29 | 23 (79.31%) | 5 (17.24%) | 5 (100.00%) | 18 (75.00%) |
| **13** | 35 | 34 (97.14%) | 30 (85.71%) | 30 (100.00%) | 4 (80.00%) |
| **14** | 34 | 33 (97.06%) | 27 (79.41%) | 26 (96.30%) | 7 (100.00%) |
| **15** | 27 | 26 (96.30%) | 25 (92.59%) | 24 (96.00%) | 2 (100.00%) |
| **16** | 45 | 45 (100.00%) | 42 (93.33%) | 42 (100.00%) | 3 (100.00%) |
| **17-18** | 61 | 61 (100.00%) | 59 (96.72%) | 59 (100.00%) | 2 (100.00%) |
| **19-20** | 24 | 23 (95.83%) | 23 (95.83%) | 22 (95.65%) | 1 (100.00%) |
| **Total** | **318** | **295 (92.77%)** | **213 (66.98%)** | **210 (98.59%)** | **85 (80.95%)** |

**Table 14: The number of children in each age group, seropositivity rates based on ACE2 blocking antibodies and vaccination rates for SARS-CoV-2 in Jaffna**

| **Age Group** | **Total Number in Each Age Group** | **Seropositivity Rate By Prescence of ACE2 Blocking Antibodies N (%)** | **Number Vaccinated N (%)** | **Number Vaccinated Seropositive N (%)** | **Number Unvaccinated Seropositive N (%)** |
| --- | --- | --- | --- | --- | --- |
| **10** | 83 | 43 (51.81%) | 3 (3.61%) | 1 (33.33%) | 42 (52.50%) |
| **11** | 83 | 49 (59.04%) | 11 (13.25%) | 9 (81.82%) | 40 (55.56%) |
| **12** | 80 | 54 (67.50%) | 14 (17.50%) | 13 (92.86%) | 41 (62.12%) |
| **13** | 95 | 88 (92.63%) | 77 (81.05%) | 74 (96.10%) | 14 (77.78%) |
| **14** | 92 | 84 (91.30%) | 77 (83.70%) | 74 (96.10%) | 10 (66.67%) |
| **15** | 91 | 79 (86.81%) | 70 (76.09%) | 67 (95.71%) | 12 (57.14%) |
| **16** | 83 | 77 (92.77%) | 71 (85.54%) | 68 (95.77%) | 9 (75.00%) |
| **17-18** | 129 | 126 (97.67%) | 120 (93.02%) | 118 (98.33%) | 8 (88.89%) |
| **19-20** | 70 | 68 (97.14%) | 68 (97.14%) | 66 (97.06%) | 2 (100.00%) |
| **Total** | **806** | **668 (82.88%)** | **511 (63.40%)** | **490 (95.89%)** | **178 (60.34%)** |

**Table 15: The number of children in each age group, seropositivity rates based on ACE2 blocking antibodies and vaccination rates for SARS-CoV-2 in Kurunegala**

| **Age Group** | **Total Number in Each Age Group** | **Seropositivity Rate By Prescence of ACE2 Blocking Antibodies N (%)** | **Number Vaccinated N (%)** | **Number Vaccinated Seropositive N (%)** | **Number Unvaccinated Seropositive N (%)** |
| --- | --- | --- | --- | --- | --- |
| **10** | 34 | 18 (52.94%) | 1 (2.94%) | 1 (100.00%) | 17 (51.52%) |
| **11** | 79 | 49 (62.03%) | 3 (3.80%) | 3 (100.00%) | 46 (60.53%) |
| **12** | 66 | 44 (66.67%) | 14 (21.21%) | 10 (71.43%) | 34 (65.38%) |
| **13** | 74 | 72 (97.30%) | 60 (81.08%) | 58 (96.67%) | 14 (100.00%) |
| **14** | 76 | 75 (98.68%) | 66 (86.84%) | 66 (100.00%) | 9 (90.00%) |
| **15** | 84 | 80 (95.24%) | 71 (84.52%) | 68 (95.77%) | 12 (92.31%) |
| **16** | 100 | 96 (96.00%) | 91 (91.00%) | 87 (95.60%) | 9 (100.00%) |
| **17-18** | 89 | 88 (98.88%) | 85 (95.51%) | 84 (98.82%) | 4 (100.00%) |
| **19-20** | 73 | 70 (95.89%) | 64 (87.67%) | 62 (96.88%) | 8 (88.89%) |
| **Total** | **675** | **592 (87.70%)** | **455 (67.41%)** | **439 (96.48%)** | **153 (69.55%)** |

**Table 16: The number of children in each age group, seropositivity rates based on ACE2 blocking antibodies and vaccination rates for SARS-CoV-2 in Kandy**

| **Age Group** | **Total Number in Each Age Group** | **Seropositivity Rate By Prescence of ACE2 Blocking Antibodies N (%)** | **Number Vaccinated N (%)** | **Number Vaccinated Seropositive N (%)** | **Number Unvaccinated Seropositive N (%)** |
| --- | --- | --- | --- | --- | --- |
| **10** | 120 | 66 (55.00%) | 1 (0.83%) | 1 (100.00%) | 65 (54.62%) |
| **11** | 138 | 85 (61.59%) | 1 (0.72%) | 0 (0.00%) | 85 (62.04%) |
| **12** | 144 | 103 (71.53%) | 14 (9.72%) | 13 (92.86%) | 90 (69.23%) |
| **13** | 155 | 137 (88.39%) | 101 (65.16%) | 95 (94.06%) | 42 (77.78%) |
| **14** | 148 | 132 (89.19%) | 104 (70.27%) | 101 (97.12%) | 31 (70.45%) |
| **15** | 156 | 132 (84.62%) | 110 (70.51%) | 100 (90.91%) | 32 (69.57%) |
| **16** | 143 | 122 (85.31%) | 97 (67.83%) | 94 (96.91%) | 28 (60.87%) |
| **17-18** | 243 | 236 (97.12%) | 227 (93.42%) | 224 (98.68%) | 12 (75.00%) |
| **19-20** | 79 | 78 (98.73%) | 76 (96.20%) | 76 (100.00%) | 2 (66.67%) |
| **Total** | **1326** | **1091 (82.28%)** | **731 (55.13%)** | **704 (96.31%)** | **387 (65.04%)** |

**Table 17: The number of children in each age group, seropositivity rates based on ACE2 blocking antibodies and vaccination rates for SARS-CoV-2 in Gampaha**

| **Age Group** | **Total Number in Each Age Group** | **Seropositivity Rate By Prescence of ACE2 Blocking Antibodies N (%)** | **Number Vaccinated N (%)** | **Number Vaccinated Seropositive N (%)** | **Number Unvaccinated Seropositive N (%)** |
| --- | --- | --- | --- | --- | --- |
| **10** | 58 | 24 (41.38%) | 4 (6.90%) | 2 (50.00%) | 22 (40.74%) |
| **11** | 53 | 40 (75.47%) | 0 (0.00%) | 0 | 40 (75.47%) |
| **12** | 56 | 40 (71.43%) | 11 (19.64%) | 9 (81.82%) | 31 (68.89%) |
| **13** | 45 | 42 (93.33%) | 42 (93.33%) | 39 (92.86%) | 3 (100.00%) |
| **14** | 63 | 59 (93.65%) | 61 (96.83%) | 57 (93.44%) | 2 (100.00%) |
| **15** | 64 | 60 (93.75%) | 56 (87.50%) | 54 (96.43%) | 6 (75.00%) |
| **16** | 49 | 47 (95.92%) | 49 (100.00%) | 47 (95.92%) | 0 |
| **17-18** | 82 | 79 (96.34%) | 77 (93.90%) | 74 (96.10%) | 5 (100.00%) |
| **19-20** | 22 | 22 (100.00%) | 20 (90.91%) | 20 (100.00%) | 2 (100.00%) |
| **Total** | **492** | **413 (83.94%)** | **320 (65.04%)** | **302 (94.38%)** | **111 (64.53%)** |

**Table 18: The number of children in each age group, seropositivity rates based on ACE2 blocking antibodies and vaccination rates for SARS-CoV-2 in Badulla**
